# Development and large-scale validation of a highly accurate SARS-COV-2 serological test using regular test strips for autonomous and affordable finger-prick sample collection, transportation, and storage

**DOI:** 10.1101/2020.07.13.20152884

**Authors:** Renata G. F. Alvim, Tulio M. Lima, Danielle A. S. Rodrigues, Federico F. Marsili, Vicente B. T. Bozza, Luiza M. Higa, Fabio L. Monteiro, Daniel P. B. Abreu, Isabela C. Leitão, Renato S. Carvalho, Rafael M. Galliez, Terezinha M. P. P. Castineiras, Leonardo H. Travassos, Alberto Nobrega, Amilcar Tanuri, Orlando C. Ferreira, Leda R. Castilho, André M. Vale

**Author notes:** **Corresponding authors:** André M. Vale and Leda R. Castilho. these authors have contributed equally (Renata G. F. Alvim, Tulio M. Lima, Danielle A. S. Rodrigues). joint senior authors (Leda R. Castilho, André M. Vale).

## Abstract

Accurate serological tests are essential tools to allow adequate monitoring and control of COVID-19 spread. Production of a low-cost and high-quality recombinant viral antigen can enable the development of reliable and affordable serological assays, which are urgently needed to facilitate epidemiological surveillance studies in low-income economies. Trimeric SARS-COV-2 spike (S) protein was produced in serum-free, suspension-adapted HEK293 cells. Highly purified S protein was used to develop an ELISA, named S-UFRJ test. It was standardized to work with different types of samples: (i) plasma or serum from venous blood samples; (ii) eluates from dried blood spots (DBS) obtained by collecting blood drops from a finger prick. We developed a cost-effective, scalable technology to produce S protein based on its stable expression in HEK293 cells. Using this recombinant antigen, we presented a workflow for test development in the setting of a pandemic, starting from limited amounts of samples up to reaching final validation with hundreds of samples. Test specificity was determined to be 98.6%, whereas sensitivity was 95% for samples collected 11 or more days after symptoms onset. A ROC analysis allowed optimizing the cut-off and confirming the high accuracy of the test. Endpoint titers were shown to correlate with virus neutralization assessed as PRNT90. There was excellent agreement between plasma and DBS samples, significantly simplifying sample collection, storing, and shipping. An overall cost estimate revealed that the final retail price could be in the range of one US dollar. The S-UFRJ assay developed herein meets the quality requirements of high sensitivity and specificity. The low cost and the use of mailable DBS samples allow for serological surveillance and follow-up of SARS-CoV-2 vaccination of populations regardless of geographical and socio-economic aspects. We hope the detailed guidelines for the development of an affordable and accurate anti-spike SARS-COV-2 ELISA, such as S-UFRJ described here, will stimulate governmental and non-governmental health agencies in other countries to engage in much-needed large-scale studies monitoring the spread and immunity to SARS-COV-2 infection.

## Introduction

The COVID-19 pandemic brought an unprecedented global health crisis. The high transmissibility of SARS-COV-2 caused an avalanche of cases within a short period of time, having caused approximately more than 130 million cases and 2,8 million deaths as of March 2021. In many cities, the number of infected individuals needing medical treatment exceeded hospitals capacity, and healthcare personnel became overwhelmed by the excess of working hours and by adverse mental health outcomes. In addition to this brutal scenario, the elevated human-to-human contagious rate obliged a severe restriction of social life including, in some cases, complete lockdown except for essential activities. As a result, this situation led to economic disruption in many countries experiencing several months of partial or complete lockdown. If easing of restrictive measures is unavoidable, it cannot be done without rigorous planning and tight follow-up. Thus, epidemiological surveillance studies are imperative to measure the spread of the disease and to estimate seroprevalence, guiding policy decisions regarding the need to keep or ease social distance measures and to maintain or not other preventive measures.

Accurate serological assays are essential tools for epidemiological surveillance studies to monitor and control epidemics. The structural spike (S) protein of SARS-COV-2 is known to be a major target for neutralizing antibodies, thus making it a key antigen for the development of specific and sensitive sero-diagnostic tests. S protein contains the receptor binding domain in its S1 subunit and is also responsible for fusion to the cell membrane through its S2 subunit^1^. Enzyme linked immunosorbent assays (ELISA) based on the S protein have been developed, showing minimal cross-reactivity with sera against circulating “common cold” coronaviruses (but some degree of cross reactivity with SARS and MERS-COV sera)^2^ and providing correlation to virus neutralizing activity^3^. However, the current unavailability of low-cost and high-quality recombinant S protein is a bottleneck for the development of affordable and reliable serological tests urgently needed by public health agencies, especially in low-income economies, to deal with the pandemic. Here, we focused on establishing the hallmarks for a low-cost ELISA based on S protein immunoreactivity, suited to be employed in epidemiological surveillance studies, evaluation of vaccine immunogenicity, with special relevance for remote regions and low-income countries with limited clinical laboratory network. Without compromising the performance of the assay, costs were cut mainly by optimizing antigen production and by simplifying sample collection and processing. Blood collection by finger prick, using dried blood spots (DBS), greatly facilitated collection, maintenance, shipping and processing of samples. The overall cost of the assay was estimated to be approximately one dollar per test.

## Methods

### Stable cell line generation

HEK293-3F6 cells (NRC Canada) growing in suspension in the chemically-defined, animal component-free HEK-TF (Xell AG) culture medium were stably transfected by lipofection using a broad-spectrum reagent (Lipofectamine 3000, Thermo Fisher) as described previously^4^. A total DNA concentration of 0.9 µg/mL was used, combining two vectors: the pαH vector (at 0.75 µg/mL) containing the sequence encoding the ectodomain (aminoacids 1-1208) of the spike protein in the prefusion conformation^1^ (kindly provided by B. Graham, VRC/NIH, also available from BEI Resources under #NR-52563), and an empty vector containing the neomycin phosphotransferase gene (at 0.15 µg/mL). Cells were maintained under selection pressure with 100 µg/mL G-418 sulfate (Thermo Fisher) from 2 days post-transfection on. 24 days post-transfection cell viability had recovered to over 90% viability, and a cell bank was cryopreserved. All steps and materials used during recombinant cell line generation, expansion and cell banking were documented in detail.

### Cell cultivation

After recovery from thawing, stably transfected cells were transferred to the chemically-defined, animal component-free HEK-GM culture medium (Xell AG) and maintained at 37°C and 5% CO_2_ under orbital agitation (180 rpm, shaker with 5-cm stroke) in vented Erlenmeyer flasks containing up to 60% of their nominal volume. In the experiments carried out in fed-batch mode, the culture medium was supplemented with a concentrated solution of nutrients (HEK-FS, Xell AG) according to the manufacturer’s instructions. Bioreactor runs were carried out in a 1.5-L stirred-tank bioreactor (EZControl, Applikon) at setpoints of pH, temperature and dissolved oxygen of 7.1, 37°C and 40% of air saturation, respectively. Cell concentration and viability were determined by trypan blue exclusion using an automated cell counter (Vi-Cell, Beckman Coulter), whereas glucose and lactate concentrations were monitored using a metabolite analyzer (YSI 2700, Yellow Springs Instruments). Presence of S protein in the supernatants was determined by spot blots: 3 µL of each sample was applied to nitrocellulose membranes, serum of SARS-COV-2 convalescent patients (1:1000) was used as primary antibody, followed by incubation with anti-human IgG(Fc) HRP-labeled antibody (Sigma, #SAB3701282) and finally addition of chemiluminescent ECL reagent (BioRad). Images were captured using a FluorChem E system (ProteinSimple), setting exposure time to be automatically determined based on the samples with strongest signal.

### S protein concentration/purification

Cell suspension harvested from cell cultures was clarified by filtration using 0.45-µm PVDF membranes (Merck Millipore) and used to perform either ultrafiltration/diafiltration (UF/DF) or affinity chromatography (AC) experiments. Concentration/diafiltration (90-fold concentration by volume) was achieved by UF/DF using centrifugal devices based on cellulose membranes with 100-kDa cut-off (Merck Millipore). Affinity chromatography was carried out using a 5-mL StrepTrap XT affinity chromatography column (Cytiva) following manufacturer’s instructions. Protein concentration, purity and identity in the eluted fractions were confirmed by NanoDrop (Thermo Fisher), silver-stained SDS-PAGE and Western blot analyses, respectively. For Western blots, serum of SARS-COV-2 convalescent patients (1:1000) was used as primary antibody, followed by incubation with anti-human IgG (Fc) HRP-labeled antibody (Sigma, #SAB3701282) and then chemiluminescent ECL reagent (BioRad).

### S-UFRJ ELISA for anti-S IgG detection

High binding ELISA plates (Corning) were coated with 50 μL of SARS-COV-2 S protein in PBS (Gibco) at 4 µg/mL concentration (unless otherwise stated in the Figures) and incubated overnight at room temperature (RT). The coating solution was removed, and 150 μL of PBS 1% BSA (blocking solution) was added to the plate and incubated at RT for 1-2 hours. The blocking solution was removed, and 50 μL of serum or plasma diluted 1:40 in PBS 1% BSA, and serially three-fold diluted, was added to the plate and incubated at RT for 2 hours. For ELISA using dried blood spots, at first circles cut from filter paper using commercially available punching devices, or single filter paper pads cut from plastic strips were used to prepare eluates by incubating for 1 hour at RT in 100 µL (filter paper circles) or 200 µL (pads) of PBS 1% BSA. In the same way as diluted serum or diluted plasma, 50 µL of eluate samples were added to the plate and incubated at RT for 2 hours. The plate was then washed five times with 150 μL of PBS. Next, 50 μL of 1:8000 goat anti-human IgG (Fc) HRP-labeled antibody (Sigma, #SAB3701282) was added to the plate and incubated for 1.5 hours at RT. The plate was then washed five times with 150 μL of PBS. At the end, the plate was developed with TMB (3,3’,5,5;-tetramethylbenzidine) (Thermo Fisher). The reaction was stopped with 50 µL of 1 N HCl, and the optical density (O.D.) was read at 450 nm with 655 nm background compensation in a microplate reader (BioRad). Results are expressed during test development either as O.D. or O.D. summation^5^, and later during application of the final test protocol as the ratio of O.D. of the sample to the cut-off. Cut-off after optimization by means of the receiver operator characteristic (ROC) analysis was defined as the sum of O.D. mean of negative controls in the same plate plus 3 times the O.D. standard deviation of negative controls.

### Commercial tests used for comparison to S-UFRJ test

A commercial ELISA to detect anti-SARS-COV-2 IgG produced by Euroimmun (#EI 2606-9601 G) and a commercial rapid diagnostic test (RDT) with separate bands for IgM and IgG detection were used following manufacters’
s instructions. The RDT is a immunochromatographic test manufactured by Hangzhou Biotest Biotech Co. and commercialized in several countries/regions, such as Brazil, Europe, USA and Australia. In Brazil, the brand name of the RDT is MedTeste (Medlevensohn).

### Sample collection from human subjects

Samples collected at the State Hematology Institute Hemorio followed a protocol approved by the local ethics committee (CEP Hemorio; approval #4008095). Samples collected at UFRJ COVID Screening and Diagnostic Center followed a protocol approved by the national ethics committee (CONEP, Brazil; protocol #30161620000005257; approval #3953368): subjects were initially interviewed and, if they accepted to participate, they signed the informed consent, answered a questionnaire (addressing demographic data, onset and type of symptoms, history of travel abroad, among other information) and had blood (venous blood and/or finger prick) and nasopharyngeal swab collected. Only symptomatic subjects who presented at least two of the following symptoms were included: loss of taste or smell, fever, shortness of breath, diarrhea, headache, extreme tiredness, dry cough, sore throat, runny or stuffy nose, or muscle aches. The protocol approved by the national ethics committee also covers the collection of samples from vaccinated individuals that had no history of SARS-CoV-2 infection to study the performance of S-UFRJ assay for evaluation of vaccine immunogenicity. Dried blood spots (DBS) were obtained by finger pricking with commercially available sterile lancets and lancing devices. Either plastic strips containing pads of filter paper, or 2.5 cm (W) x 7.5 cm (L) filter paper with three blood spots from the same volunteer, were used to collect whole blood from finger pricks.

### Plaque reduction neutralization test (PRNT)

In order to determine the titers of neutralizing antibodies in serum samples, sera were first heat-inactivated at 56°C for 30 min, and two-fold serial dilutions were incubated with 100 PFU of SARS-COV-2 (GenBank #MT126808.1) for 1 hour at 37°C to enable neutralization to occur. Virus-serum mixture was inoculated into confluent monolayers of Vero cells seeded in 12-well tissue culture plates. After 1 hour, inoculum was removed and a semisolid medium (1.25% carboxymethylcellulose in alpha-MEM supplemented with 1% fetal bovine serum) was added. Cells were further incubated for 72 hours and then fixed with 4% formaldehyde solution. Viral plaques were visualized after staining with crystal violet dye solution. PRNT end-point titers were expressed as the reciprocal of the highest serum dilution for which the virus infectivity is reduced by ≥90% (PRNT_90_) when compared with the average plaque count of the virus control. All work involving infectious SARS-COV-2 was performed in a biosafety level 3 (BSL-3) containment laboratory.

## Results

### Antigen expression, production and purification

For recombinant production and straightforward purification of a heavily glycosylated protein such as the SARS-COV-2 spike protein, mammalian cell culture in serum-free media is the option of choice. We expressed the soluble ectodomain of the spike (S) protein in a stabilized prefusion conformation^1^ in serum-free, suspension-adapted cell cultures. Initial transient transfections indicated that expression levels evaluated on days 2 and 4 post-transfection were higher in HEK293 cells than in CHO-K1 cells (Fig. 1A, left panel). However, differently from previous studies that adopt transient protein expression techniques^3^, we focused on stable and constitutive gene expression because transgene integration in the cell genome enhances scalability and significantly decreases costs of recombinant protein production in mammalian cells^4^. Due to the challenges posed by the pandemic, such as the urgency and the disruption of international supply chains for reagents and synthetic gene constructs, we decreased time and costs by using an old-fashioned technique of co-transfecting the plasmid containing the S gene with an intellectual property-free plasmid containing an eukaryotic selection marker. A stable recombinant HEK293 cell line showing higher expression than transiently transfected HEK293 cells was generated (Fig. 1A, center panel) and banked 24 days post-transfection. Stability of S protein expression by this cell line has been proven so far for up to 100 days post-transfection (Fig. 1A, right panel). Hence, confirmation of expression stability is required for developing cost-saving, long-lasting batch-refeed or perfusion technologies for cell culture.

**Figure 1:**
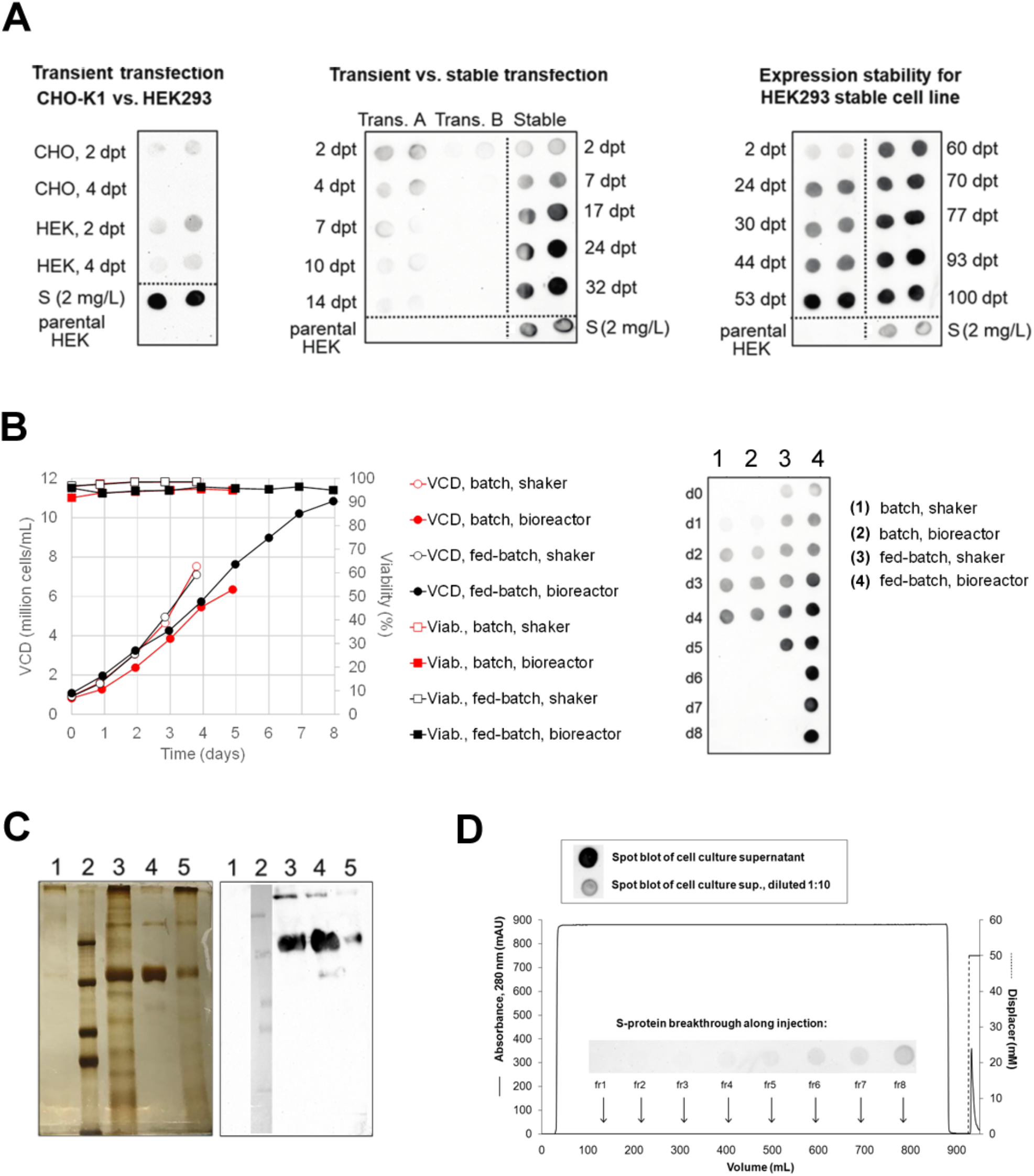
S protein production by stable recombinant HEK293 cells and its purification from cell culture supernatant. **(A)** Left panel: CHO-K1 and HEK293-3F6 were transiently transfected with plasmid DNA containing the S protein gene. Center panel: due to the higher levels of protein secreted by HEK293 cells, this cell line was used in transient transfections at high (H, 2 µg/mL of pαH plasmid) and low (L, 0.75 µg/mL of pαH plasmid) DNA concentration, as well as in a stable transfection that comprised co-transfection of a second vector containing the neo selection marker. A well documented cell bank was cryopreserved on day 24 post-transfection. Right panel: stability of expression of the secreted protein has been confirmed so far for up to 100 days post-transfection. **(B)** Left panel: high cell viabilities and viable cell densities (VCD) were achieved in the cultivation of the stable cell line in shake flasks and 1.5-L stirred-tank bioreactors, in batch and fed-batch mode, using chemically-defined, animal component-free media. Right panel: spot blot for detection of S protein in the cell culture supernatant on different days of culture, showing high titers of secreted S protein. **(C)** S protein identity was confirmed by Western blot analysis, confirming the molecular mass of approximately 180 kDa. SDS-PAGE showed a rather low purity for the concentrated/diafiltered (UF/DF) sample, but a very high purity for the sample purified by affinity chromatography (AC). 1: Cell culture supernatant from non-transfected parental HEK293-3F6 cells. 2: Molecular-mass standard containing proteins at 250, 150, 100 and 75 kDa. 3: UF/DF sample after 90-fold volumetric concentration and buffer exchange into PBS. 4. Eluate from affinity chromatography. 5. Supernatant from stable recombinant cell line. **(D)** A typical chromatogram of the affinity chromatography purification. Fractions of the flow-through (fr1 to fr8) were collected during sample injection and analyzed by spot blot to evaluate progressive saturation of affinity ligands, showing a high binding capacity and a low degree of S protein leakage. For comparison, spot blots of the injected cell culture supernatant, or a 1:10 dilution, are shown in the box above the chromatogram. In all immunoblots, presence of S protein was detected using a pooled serum of SARS-COV-2 convalescent patients (1:1000) as primary antibody, followed by incubation with anti-human IgG-HRP and chemiluminescent detection.

In order to decrease costs and logistics burden, cell culture media available as dried powdered media were selected to be tested both in shake flasks and stirred-tank bioreactors at 300-mL and 1.5-L scale, respectively. The chosen medium (HEK-GM) was able to provide robust cell growth and efficient recombinant protein production (Fig. 1B). Carrying out cell culture in fed-batch mode by adding pulses of a concentrated nutrient solution over cell cultivation time avoided nutrient depletion and significantly increased viable cell density and secreted S protein levels, allowing enhanced production of the recombinant protein (Fig. 1B, right panel). Protein isolation from cell culture supernatant was investigated by ultrafiltration/diafiltration (UF/DF) and affinity chromatography (AC) techniques. In spite of the large S protein size (∼170 kDa) and the use of a 100-kDa cut-off membrane device, UF/DF was not able to efficiently remove all smaller contaminating proteins, and an AC resin bearing a streptavidin mutein ligand was used to obtain the protein in high purity (Fig. 1C). The affinity resin has shown a relatively high dynamic binding capacity for the S protein (Fig. 1D), and has so far been used for 50+ adsorption/elution/regeneration cycles with no detectable decrease in performance, reducing its impact on the final costs of the purified protein.

### Early development of the assay from a limited number of samples

Early on in the pandemic in Brazil, March / April 2020, development of the test was started by using a relatively limited number of samples that were available by then to compare S protein preparations of lower (UF/DF) or higher purity (AC) for the detection of anti-S antibodies in plasma samples. Nineteen positive or negative samples, collected from individuals scored as positive by PCR for SARS-COV-2 (n=16), from a healthy non-infected individual (“post-pandemic negative”, n=1) or collected before SARS-COV-2 emergence (“pre-pandemic negative”, n=2), were tested for immunoreactivity against S protein preparations obtained either by UF/DF or AC. Considering the limited number of samples available at this stage, the threshold value to discriminate between negative and positive samples was considered as the mean plus 3 standard deviations of O.D. of the three negative controls added to each plate.

Comparison of the results of the ELISAs carried out with S protein of different purities (UF/DF or AC) showed that the assay performance could be greatly improved by the use of the highly purified AC antigen, showing much better discrimination between positive samples and negative controls (Fig. 2A). For comparison, the samples were evaluated also by means of an imported rapid diagnostic test (RDT), which is approved for commercialization in many countries including Brazil. Interestingly, sample #1, which was positive by PCR, but negative for IgG by RDT, clearly scored as positive in the AC S-protein ELISA.

**Figure 2:**
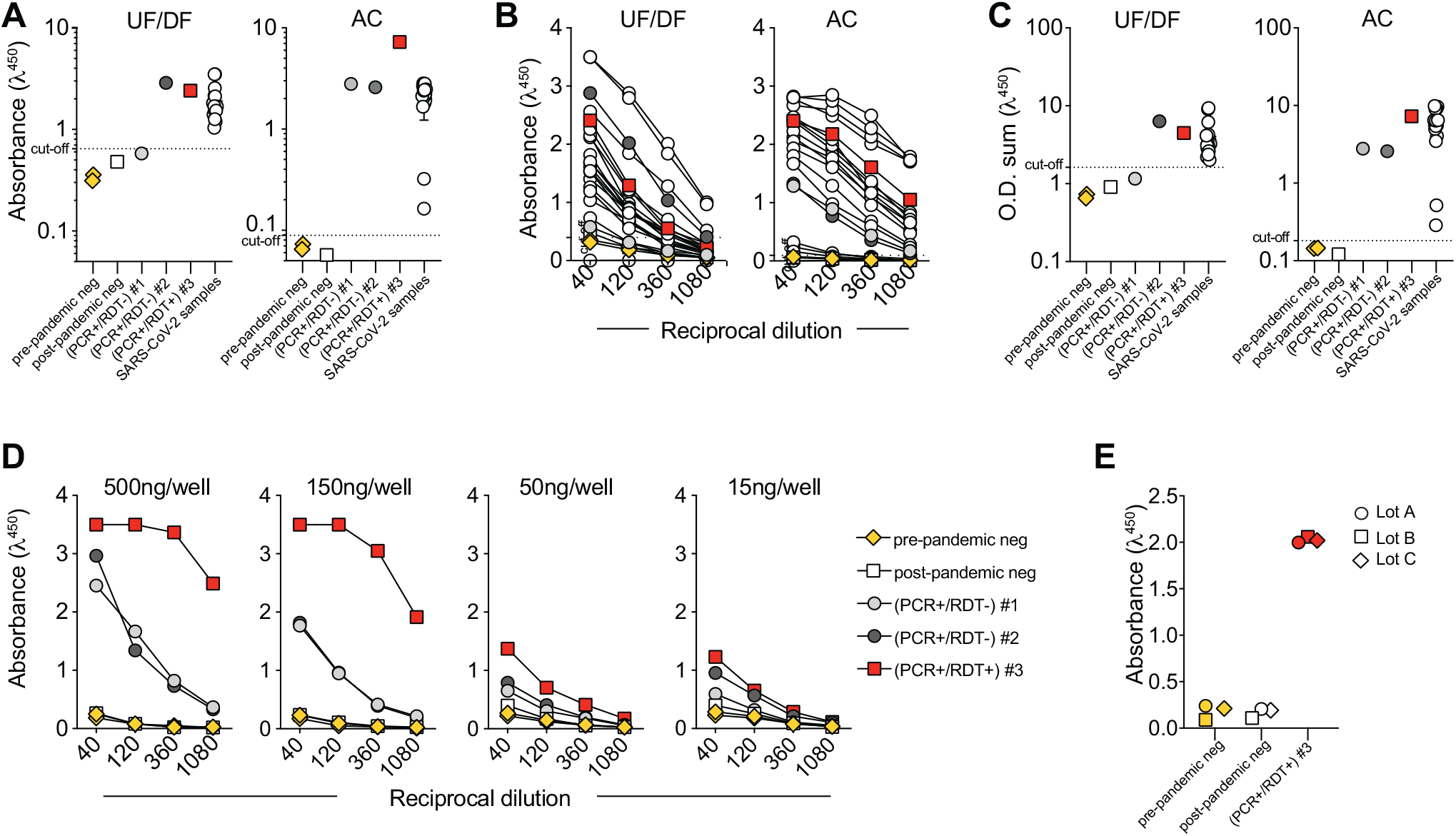
Comparison of ELISAs with UF/DF- or AC-purified S protein. **(A)** ELISA performance using low-purity (UF/DF) or high-purity (AC) S protein antigen to coat plates. A total of 22 samples were used: 15 sera from COVID-19 convalescent patients (SARS-COV-2 samples, open circle), two control samples collected until 2018 (pre-COVID-19, yellow diamond), one post-COVID-19 control sample from a healthy individual, two samples from SARS-COV-2 infected individuals characterized as PCR+/RDT-(#1 light gray filled circle, #2 dark gray filled circle), and one sample from a SARS-COV-2 convalescent patient who had the severe form of the disease (#3, red square). The cut-off for antigen evaluation was defined as [mean + 3 standard deviations (X+3SD)] of the O.D. of negative controls. **(B)** Samples used in (A) were titrated in four serial dilutions (1:40, 1:120, 1:360 and 1:1080), both for UF/DF or AC ELISA. **(C)** O.D. summation of the titration curves shown in (B); symbols as in (A). **(D)** Samples used in (A) were titrated in four serial dilutions (1:40, 1:120, 1:360 and 1:1080) for AC ELISA using different S-protein coating concentrations (0.3 – 10 µg/mL, corresponding to 15 – 500 ng/well). **(E)** Three selected samples from (D) were diluted 1:40 to evaluate antigen lot-to-lot consistency, inter- and intra-operator variability of the AC ELISA. Representative data from 3 experiments using 3 different lots of the AC-purified S protein (lots A to C) are shown. Experiment with lot A was performed by one operator, and a second operator performed experiments with lots B and C.

Better discrimination between positive and negative samples enabled by the AC antigen was also evident in titration curves (Fig. 2B), which were used to calculate the O.D. summation for each sample, evidencing again the superiority of the highly pure S protein as ELISA antigen (Fig. 2C). Using selected plasma samples from negative controls and PCR-confirmed patients, we established that a minimum of 150 ng of high-purity S protein per well (i.e. by coating wells with 50 µL of a 3 µg/mL antigen solution) enables clear discrimination of positive and negative samples (Fig. 2D). Additionally, we evaluated antigen lot-to-lot consistency, as well as inter- and intra-operator variability of the ELISA. Representative data from experiments done by different operators and using different lots of the AC-purified S protein are shown in Fig. 2E and corroborate the reliability of the recombinant antigen and of the test as a whole. The ELISA test with AC-purified S protein was named S-UFRJ test.

### Late assay development using a broader panel of samples

As broader sample panels became available, a final development of the test was performed. We applied the S-UFRJ ELISA to evaluate the presence of anti-S IgG in 210 samples, including pre-pandemic negative controls and samples from symptomatic individuals diagnosed by PCR as positive for SARS-COV-2. The PCR-positive cohort consisted of 66 samples from 38 symptomatic individuals whose blood samples were obtained at different time points after symptoms onset. Negative control samples were collected either until 2018 from healthy individuals (pre-pandemic negative controls, n=124) or in early 2020 from healthy individuals who tested negative for SARS-COV-2 by PCR (post-pandemic negative controls, n=20), comprising a total of 144 negative control samples.

Results of serological tests are usually interpreted based on the ratio of the O.D. of the given sample to a pre-established cut-off. Since at this stage of development an optimized cut-off was not yet available, we adopted a conservative rationale of classifying as undetermined all samples having an O.D. value between one and two times the mean plus standard deviation (SD) of the O.D. of negative controls in the same plate. All samples showing an O.D. below the lower threshold value (mean + 1 SD) were considered negative, and all samples having an O.D. above the higher threshold value (2*[mean + 1 SD]) were considered positive.

Out of 144 negative control samples, 142 scored as negative, revealing a very high specificity of the test of 98.6% (Fig. 3A). Interestingly, we have observed that a large fraction of these pre-pandemic negative samples display immunoreactivity against spike proteins from other “common-cold” coronaviruses, highlighting the ability of the S-UFRJ assay to discriminate immunity among different coronaviruses (data not shown).

**Figure 3:**
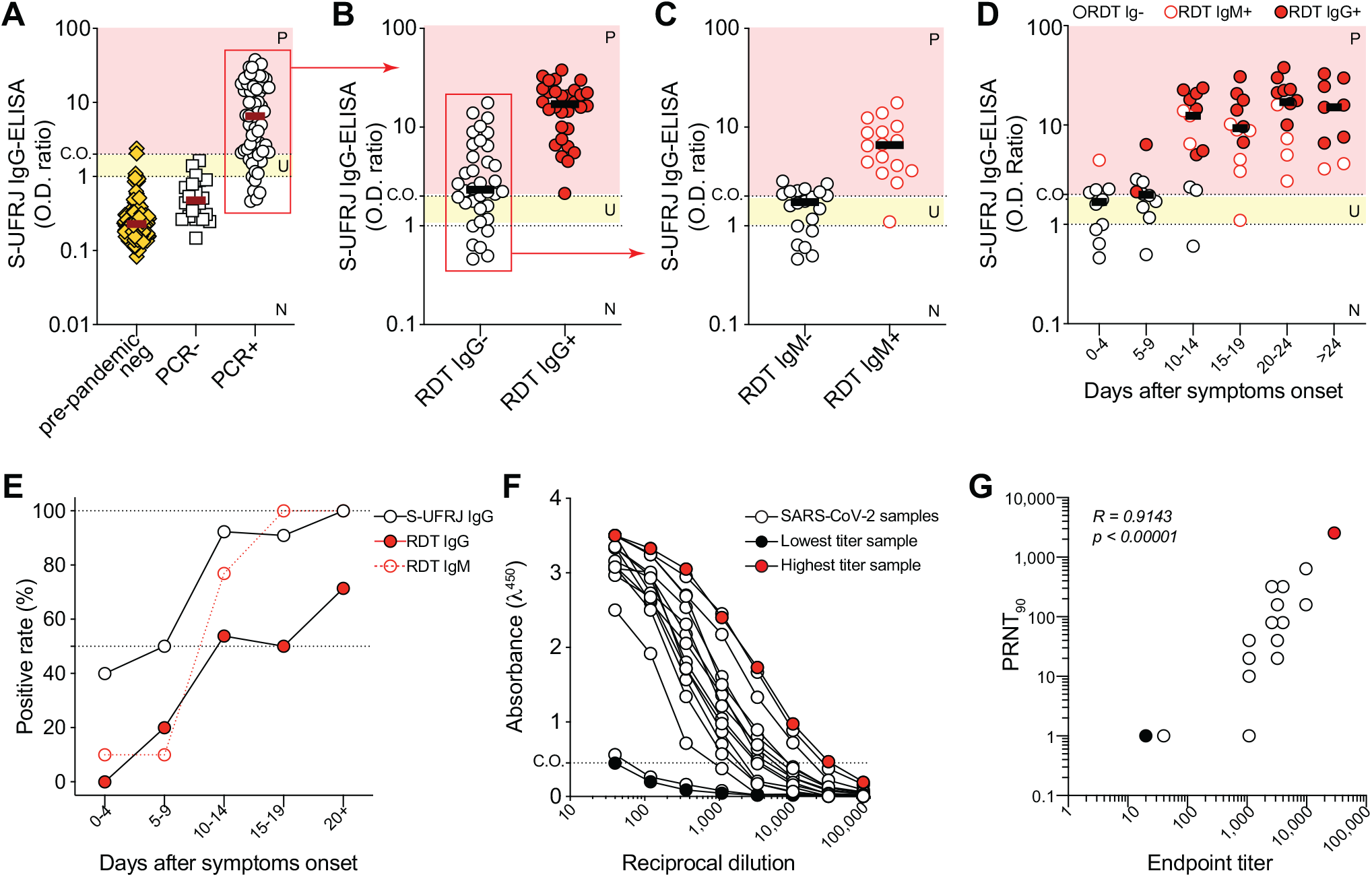
Validation of S-UFRJ ELISA, which comprises the AC-purified antigen, for early detection and quantification of anti-S IgG serum antibodies. **(A)** Anti-S IgG antibody detection in samples from healthy individuals, obtained as pre-pandemic controls (n=124, yellow diamond) or post-pandemic controls from individuals who tested negative for the virus by PCR (n=20, open square), and samples from COVID-19 patients who were PCR-positive (n=66, open circle). Relative levels of antibodies are shown as O.D. ratio of values of individual samples to the [mean + 1 standard deviation (X+1SD)] of the O.D. average of the negative controls in the same ELISA plate. Sera samples were routinely diluted at 1:40. For this stage of development of the assay, it was conservatively established that an O.D. ratio below 1 indicates a negative result (N), an O.D. ratio above 2 indicates a positive result (P), and an O.D. ratio between 1 and 2 is considered undetermined (U). **(B)** The PCR-positive samples shown in (A) were tested for anti-SARS-COV-2 IgG by a commercial rapid diagnostic test (RDT) (MedTeste, Medlevensohn, Brazil, imported from Hangzhou Biotest Biotech Co.). Samples were then grouped by IgG reactivity according to the RDT result (RDT IgG-, open circle; RDT IgG+, red filled circle), and their anti-S IgG levels measured by the S-UFRJ ELISA are plotted. **(C)** Anti-S IgG levels measured by the S-UFRJ ELISA for the RDT IgG-negative samples shown in (B), grouped according to the RDT IgM result as RDT IgM-(open circle) or RDT IgM+ (open red circle). **(D)** Levels of anti-S IgG in samples grouped according to the timepoint of sample collection given in days after symptoms onset (DASO); symbol legends are indicated in the figure. **(E)** Positive rates versus DASO for anti-S IgG measured by S-UFRJ ELISA (open circle), for IgM measured by RDT (open red circle) and for IgG measured by RDT (red filled circle). **(F)** S-UFRJ IgG titration of 16 samples from COVID-19 convalescent patients (SARS-COV-2 samples); samples with the highest and lowest endpoint titer of the group are indicated in solid red and solid black colors, respectively. **(G)** Correlation between S-UFRJ ELISA IgG endpoint titers and neutralization (PRNT_90_) for samples titrated in (F) (n=16). Three samples with PRNT_90_ values <10 are plotted as 1. Statistical analysis was performed using Pearson’s test.

Considering all samples from PCR-positive individuals, regardless of the time of collection after symptoms onset, 53 out of 66 samples (80.3%) were IgG positive (Fig. 3A). For comparison, all samples were also tested for IgG using the RDT, and only 30 out of 66 samples were positive for IgG (45.5%) (Fig. 3B). In order to gain insight into the samples from symptomatic PCR-positive individuals that scored negative for anti-S IgG in the S-UFRJ ELISA, samples were grouped as IgM- and IgM+ according to the RDT (Fig. 3C). We found that 14 out of 15 samples that scored negative for anti-S IgG in our ELISA, also scored as negative for IgM in the rapid test (Fig. 3C). This result suggests that samples from PCR+ individuals that scored negative for anti-S IgG in our ELISA, may have been collected in the very beginning of the disease, only a few days after symptoms onset (DASO), and for that reason also scored negative for IgM in the RDT. Indeed, when the results of S-UFRJ ELISA were charted against DASO, this was confirmed: samples were increasingly scored as positive for anti-S IgG according to the time point they had been collected after symptoms onset (Fig. 3D). Accordingly, PCR+ individuals that have been sequentially sampled on different DASO and scored negative in their first sampling, later converted to seropositivity for anti-S IgG (data not shown). Anti-S IgG seroconversion rate, as scored by S-UFRJ ELISA, increased progressively from 41.66% (days 0-4) to 100% (20+ days) as a function of DASO, being above 90% for all samples collected 10 or more days after symptoms onset (Fig. 3E). Of note, even for DASO of 20+ days, the RDT reached a maximum positive rate of 71.4%, whereas the S-UFRJ ELISA scored a rate of 100%. The results show the superiority of S-UFRJ ELISA when compared to the commercial RDT used for comparison, with higher sensitivity and earlier detection of seroconversion in PCR-positive symptomatic individuals.

A previous study using another S protein-based ELISA has observed a correlation between anti-S or anti-RBD (the receptor binding domain within the S protein) IgG titers and virus neutralization^3^. To address whether the S-UFRJ ELISA could also bring information about the neutralizing capacity of positive samples, we compared anti-S IgG titers and virus neutralizing titers. We tested plasma samples of COVID-19 convalescent donors for their SARS-COV-2 in vitro neutralization capacity as measured by classic plaque reduction neutralization test (PRNT), using a Brazilian SARS-COV-2 isolate. Importantly, analogously to what has been shown by Amanat and colleagues^3^, we found that the higher the anti-S IgG ELISA endpoint titers (Fig. 3F), also the higher the neutralization titers were (Fig. 3G), resulting in a high correlation between those titers (Pearson’s R=0.9143, p<0.00001). Hence, S-UFRJ ELISA also provides an important functional correlation with SARS-COV-2 neutralization capacity.

### ROC analysis and cut-off optimization

The receiver operator characteristic (ROC) analysis is a standard procedure for assessing diagnostic performance, since it allows both to evaluate accuracy with no need for a predetermined cut-off^6^ and to determine an optimized cut-off value^7^. Using a sample panel provided by the state blood bank of Rio de Janeiro (Hemorio) comprising of 420 positive and 68 pre-pandemic negative samples, a ROC analysis was performed. The ROC curve (Supplementary Fig. S1) revealed a very high accuracy of the S-UFRJ test as given by an area under the curve (AUC) approaching 1. The curve was built by determining the sensitivity and specificity of the test for a wide range of cut-off values, and allowed establishing the optimized cut-off resulting in the best combination of specificity and sensitivity. The optimized cut-off was determined to be equal to the O.D. mean of the negative controls in the same plate (also usually known as calibrators) plus 3 times the standard deviation that was previously determined for a plate full of pre-pandemic negative controls (3*0.016 = 0.048). Importantly, by establishing an optimized cut-off determined based on a large amount of samples, the definition of a realistic (instead of conservative) range for undetermined samples became possible. Thus, for final application of the S-UFRJ test using the optimized cut-off value determined herein, the recommended range for classification of samples as undetermined is a ratio of O.D. to cut-off between 0.9 and 1.1, as also used by most commercial diagnostic ELISA tests.

### Performance validation and comparison to a widely available commercial ELISA assay

To validate S-UFRJ test performance, an additional sample panel comprising of 437 positive samples from individuals who had tested positive by PCR and were followed over time, providing well-characterized samples with a wide range of days after symptoms onset (DASO of 0-98 days), was used. These samples were measured side-by-side by the S-UFRJ ELISA and by a high-reputation IgG ELISA based on the S1 subunit of the spike protein, which is commercialized worldwide by the company Euroimmun.

Side-by-side comparison of tests using the same sample panel is important, because sensitivity data can change a lot according to DASO of samples included in a given panel. We chose to use a sample panel having 17.2% of samples collected at early timepoints (DASO of 0-10 days), when seroconversion possibly had not yet occurred, in order to get a true insight into performance of both assays.

As shown in Table 1, the S-UFRJ ELISA allows earlier detection of seroconversion and presents a greater sensitivity than the Euroimmun S1-ELISA. Considering samples with DASO of 11-98 days, S-UFRJ sensitivity was 95.0% and Euroimmun sensitivity was 86.5%. If all 437 samples of the panel are included in the computation (i.e. if early samples with DASO of 0-10 days are also included), then S-UFRJ sensitivity was 82.4% and Euroimmun sensitivity was 73.7%. Importantly, the higher sensitivity of the S-UFRJ assay is not achieved at the expense of specificity, since both assays present very similar specificities (98.6% for S-UFRJ as determined herein, and 98.7% for the Euroimmun IgG ELISA according to the product package insert).

**Table 1.** Evaluation of the S-UFRJ ELISA and of an IgG ELISA commercialized by the company Euroimmun. A panel of 437 samples from PCR-positive individuals who were followed along time was used. For S-UFRJ data analysis, the final optimized assay parameters were adopted (cut-off equal to mean + 3*SD, and O.D. ratio interval of 0.9-1.1 for undetermined samples). For the Euroimmun ELISA, test was carried out according to manufacturer’s instructions.

| Days after symptoms onset (DASO) | Number of samples | S-UFRJ ELISA |  | Euroimmun ELISA (S1 protein) |  |
| --- | --- | --- | --- | --- | --- |
|  |  | Number of positive samples | Sensitivity (%) | Number of positive samples | Sensitivity (%) |
| 00-05 | 33 | 3 | 9.1 | 1 | 3.0 |
| 06-10 | 42 | 13 | 31.0 | 8 | 19.0 |
| 11-15 | 83 | 70 | 84.3 | 53 | 63.9 |
| 16-20 | 62 | 59 | 95.2 | 53 | 85.5 |
| 21-25 | 56 | 54 | 96.4 | 50 | 89.3 |
| 26-30 | 54 | 54 | 100.0 | 53 | 98.1 |
| 31-98 | 107 | 107 | 100.0 | 104 | 97.2 |
| **11-98** | **362** | **344** | **95.0** | **313** | **86.5** |
| **All (0-98)** | **437** | **360** | **82.4** | **322** | **73.7** |

### Simplification of sample collection by using eluates from dried blood spots obtained by finger prick

The cost per sample of serological assays for COVID-19 is an important aspect concerning large-scale public health actions, especially in low-income countries. ELISA samples are routinely based on venous blood collection followed by serum or plasma preparation. This requires clinical laboratory services, as well as refrigerated sample storage and transport, which would significantly increase the cost of the S-UFRJ ELISA. More relevant, it would severely limit the usage of the assay for epidemiological surveillance studies in world regions lacking an appropriate network of clinical laboratories. To overcome this critical limitation, we evaluated a simple means for storage and transport of blood samples, by collecting blood drops obtained by finger prick in filter paper, resulting in dried blood spots (DBS) that enable sampling in remote regions, or regions lacking a laboratory network. For that, the assay was further standardized to use eluates from DBS (Fig. 4A), a simple, low-cost and low-complexity method^8^. Importantly, titration curves of plasma samples, or eluates from DBS collected in filter paper, displayed comparable results (Figs. 4B and 4C). Additionally, plastic strips with one or more filter paper pads allowed further precision in sampling and storing for eventual retesting (Fig. 4D). Consistently, O.D. ratios obtained for plasma samples and DBS, either in filter paper or in pads, showed very high correlation (Figs. 4E and 4F), as well as high reproducibility between pads from a given strip (Fig. 4G). Thus, by using dried blood spots, the low cost of the test was further warranted. We have confirmed that blood samples collected in filter paper and kept in plastic bags can be preserved for at least 2 months without altering their serological result (data not shown). Importantly, the elution volume of 200µL used for DBS pads allows up to 4 experiment replicates per pad. Additionally, as each plastic strip can contain multiple pads, it also permits additional testing for immunoreactivity against other SARS-COV-2 antigens (e.g. RBD or nucleocapsid protein), or for cross-reactivity evaluation against related viruses^2^.

**Figure 4:**
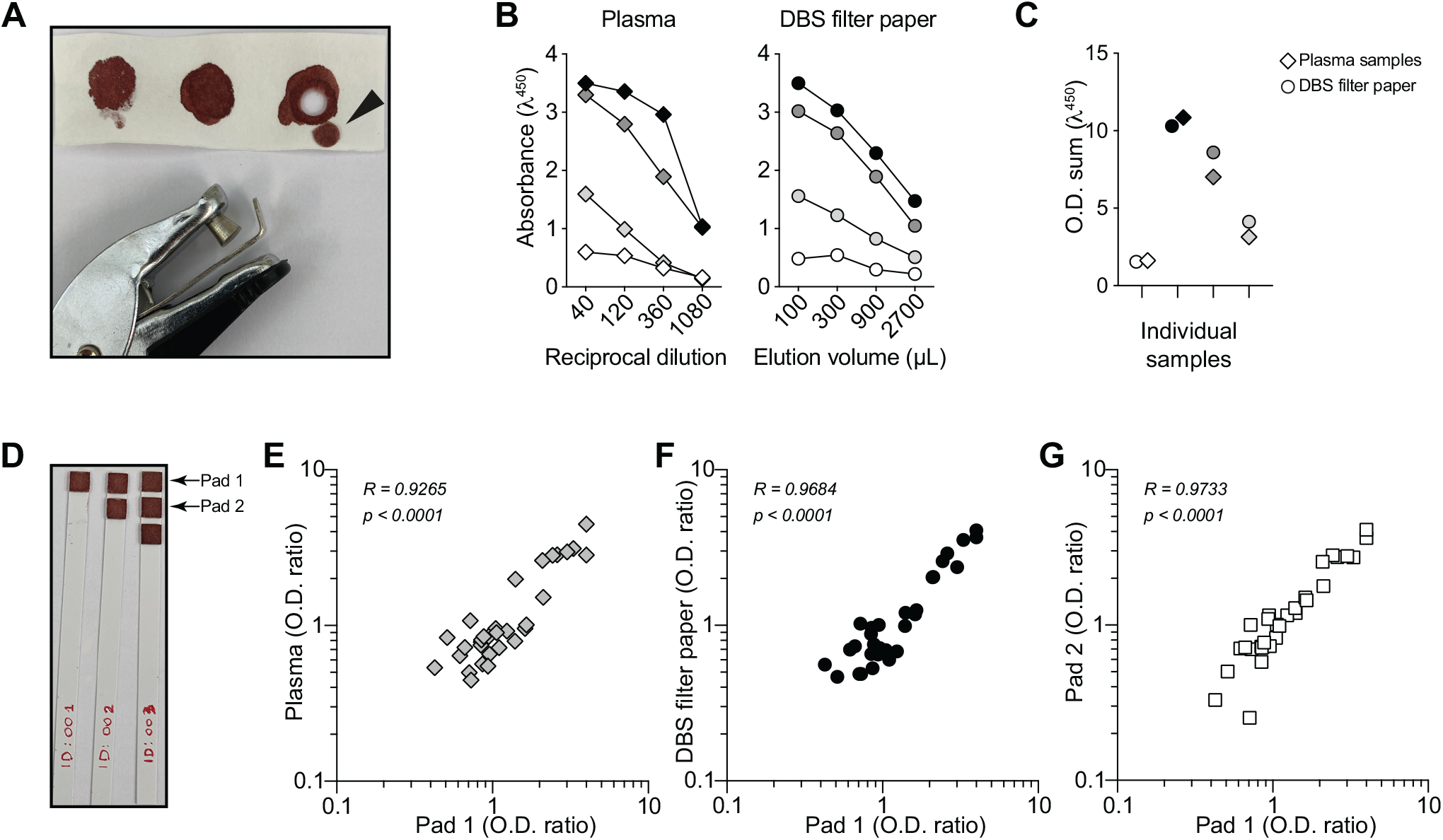
S-UFRJ test optimization for sample collection amenable for field studies. **(A)** Dried blood spots (DBS) obtained by finger pricking with commercially available lancing devices: a 2.5 cm (W) x 7.5 cm (L) filter paper with three blood spots from the same volunteer and a commercially available paper hole punching device were used to prepare a DBS disk (arrowhead) from which blood was eluted for ELISA testing. **(B)** S-UFRJ ELISA comparing the O.D. values for plasma samples in increasing serial dilutions and for the corresponding eluates prepared by incubating the DBS disks in increasing volumes of buffer. **(C)** O.D. summation of the data shown in (B). **(D)** Dried blood spots collected in plastic strips containing 1, 2 or 3 pads of filter paper. **(E-G)** correlations of O.D. ratios between samples collected in the strip pad 1 and either the respective plasma sample (E), the respective DBS in filter paper (F), or the second pad of the same strip (G). In E-G, plasma dilution was 1:40, DBS in filter paper disks were eluted in 100 µL of PBS 1% BSA, and DBS in single pads were eluted in 200 µL PBS 1% BSA. Statistical analysis was performed using Pearson’s test.

### Application of the S-UFRJ assay for evaluation of vaccine immunogenicity

Vaccination against SARS-COV-2 infection has already started in several countries including Brazil. Multiple vaccine preparations using different antigenic preparations and adjuvants are currently being employed. Phase 3 clinical studies have reported variable effectiveness depending not only on the vaccine preparation but also on population being addressed. In Brazil, as of march 2021, two different vaccine preparations are being administered: Coronavac and Astra/Zeneca. The Coronavac vaccine is based on antigenic preparations from inactivated SARS-COV-2 purified after replication in VERO cells, in vitro; the substance used as adjuvant is aluminium hydroxide. Compared to other vaccines, such as those produced by Pfizer, Moderna, Astra/Zeneca and the Sputnik V by the Gamaleya Institute, the effectiveness of Coronavac was the lowest ^15-18^. Therefore, the real-time monitoring of the immunogenicity of vaccination with Coronavac, shortly after administration of the first and second doses is a key information for the prognostic of the real impact of this vaccine in the population.

Here we had the opportunity to obtain serum samples from a small cohort of 15 individuals that have no history of SARS-COV-2 infection and were vaccinated with Coronavac. We have tested these samples for Ig seroconversion using the S-UFRJ ELISA. For each individual, the surveyal study contains samples collected on different time points, before and after the first and second doses of the vaccine. The results revealed that 11 out of 15 individuals developed IgG titers anti-S above the cut-off after the first dose of the vaccine. Of note, all individuals developed IgG titers anti-S after the administration of the second dose (Fig. 5). It is interesting to note that the IgG anti-S titers after vaccination are comparable with the titers presented by asymptomatic patients that recovered from natural infection (Fig. 4E).

**Figure 5:**
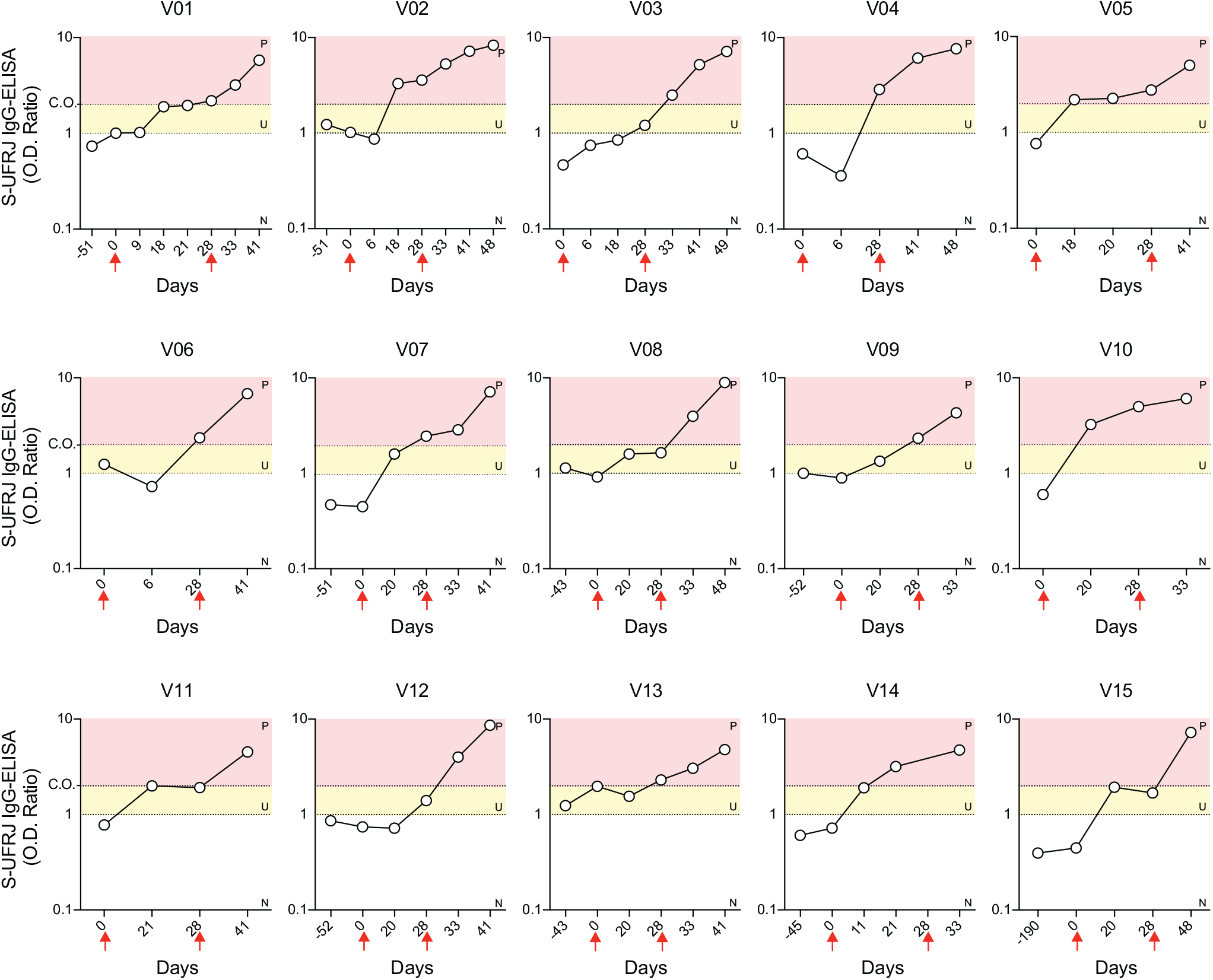
S-UFRJ test applied to periodic monitoring of anti-S IgG antibodies following vaccination. Dried blood spots were collected in plastic strips containing three pads of filter paper. For each vaccinated individual (V01 to V15), DBS were collected on different time points, before and after the first and second doses of the vaccine, indicated by the red arrows.

### S-UFRJ test costs

Taking into account the simplicity of the DBS method, the overall cost of the S-UFRJ test is significantly decreased. In terms of raw materials (consumables) needed for collecting samples and performing the assay, a detailed calculation revealed that consumables costs vary between two and four USD dimes per test. Two scenarios were considered (Supplementary Table S1): one scenario considering items sold by international vendors in the country, and the second one considering items produced locally and adopting other saving measures that we confirmed not to compromise the performance of the test (data not shown). Based on techniques routinely employed in economic feasibility studies^9^, shown in Supplementary Table S2, the final price of the test was estimated by including in the computation other manufacturing costs (that include labor, utilities, laboratory charges, insurance, among others) and general expenses (which account for administrative, research, development, distribution and marketing costs). Costs can vary significantly from country to country, but we expect that the two scenarios in Tables S1 and S2 can represent most realities reasonably well. The final retail price estimated for the S-UFRJ test from sample collection to test result is approximately USD 1 per single-well test of the sample.

## Discussion

Serological tests for the detection of anti-SARS-COV-2 antibodies are instrumental for epidemiological surveillance studies in the present COVID-19 global health crisis. Assessing the spread of SARS-COV-2 with the help of serological assays provides the data input needed for policy decisions about social distancing and other preventive measures. As discussed for the case of Spain^10^, population-based epidemiological studies are a key element to inform authorities about the need for maintaining public health measures to avoid new epidemic waves, thus allowing scientifically driven decision making by governments.

Reliable epidemiological data require accurate diagnostic tests. A systematic review and meta-analysis on serological tests for COVID-19 showed that the evidence on the accuracy of existing point-of-care lateral flow tests (rapid diagnostic tests - RDT) is particularly weak and does not support their continued use^11^. Traditional ELISA tests and chemiluminescent assays showed better performance^11^, but depend on collecting venous blood to obtain serum or plasma, thus being demanding in regard to sample collection, processing, storage and transport. Of note, both these types of tests are expensive, with retail prices in the range of $50 to $150 being common even in lower income countries as Brazil. In this context, S-UFRJ ELISA stands out as an instrumental tool for epidemiological surveillance studies. It combines low cost and simple sample collection by finger prick/DBS, with the important advantage of a high accuracy, given by a specificity of 98.6% and sensitivity of 95.0% for samples collected 11 or more days after symptoms onset. The quality of the assay is warranted by the use of the spike protein, which is less conserved in coronaviruses and thus less prone to induce cross-reactivity^12-13^. Moreover, the recombinant, highly purified spike protein used herein consists of the full-length ectodomain and is produced in the prefusion trimeric conformation, thus maintaining the most neutralization-sensitive epitopes^1^ and reliably mimicking the antigenic properties of the virus upon infection and spread in the organism.

Serological surveillance studies are urgently needed for many reasons, including to establish priorities for vaccination policies and evaluation of vaccine immunogenicity. In spite of more vaccines being manufactured and approved for human use, it is expected that a large percentage of the world population may wait long before being vaccinated because of insufficient industry production. Thus, monitoring seroprevalence will still remain highly relevant for many years. Considering that most vaccines under development are based on the spike protein (administered either as protein itself or as a nucleic acid or viral vector encoding it for in vivo expression), in the future discrimination between immunity resulting from infection or vaccination could in principle be assessed by using the DBS eluate volume to perform in parallel the S-UFRJ test and another serological assay using a different viral antigen not present in the vaccine(s) to be approved. Furthermore, it is important to note that immunity though vaccination may not be long lasting, therefore periodic monitoring of anti-S IgG will be necessary to evaluate the decline of humoral protection and the need for a new round of vaccination.

Rapid diagnostic tests are currently in widespread usage around the globe. However, beside their poor accuracy, their high cost pose a major obstacle for large-scale epidemiological surveillance, rendering these studies virtually unfeasible in less wealthy nations. In this regard, the S-UFRJ ELISA test presented a much superior performance when compared to a RDT employed for comparison, which is broadly commercialized in Brazil and also globally available. S-UFRJ also showed similar or even better sensitivity when compared to a high-reputation ELISA test produced in Europe and available globally. Importantly, the S-UFRJ test has an estimated cost of just one US dollar per single-well evaluation of samples. This is an essential feature of the assay: it can not only enable broad and reliable serological surveillance in populations, regardless of their geographical and socio-economic aspects, but can also allow governments to save very significant amounts of financial resources that could be invested in other types of countermeasures to fight the health and socio-economic consequences of COVID-19 pandemic^14^, with special relevance for low-income countries. The S-UFRJ assay is being currently used in Brazil in a large-scale serological study involving 10,000 individuals in the state of Rio Grande do Sul (Epicovid-RS)^19^. This study was only possible to be done because of the low costs of the assay. We hope the detailed guidelines for the development of an affordable and accurate anti-spike SARS-COV-2 ELISA, such as S-UFRJ described here, will stimulate governments and non-governmental health agencies in other countries to engage in similar large scale studies monitoring the spread and immunity to SARS-COV-2 infection.

## Data Availability

The raw data of all experiments are available upon request.

## Contributors

AMV, LRC and OCFJ had the idea for the study and contributed to study design. RGFA, TML, DASR, DPBA, OCFJ and AMV were involved in the experiments related to antigen production, assay development and validation. FFM, VBTB, RSC and LHT gave practical laboratory support and performed analyses. LMH and FLM performed the virus neutralization assays. ICL, RMG, TMPPC, OCFJ and AT contributed to sample collection and characterization. LHT, AN, OCFJ, AMV and LRC contributed to data analysis and interpretation. AN, AMV and LRC wrote the manuscript. LMH, LHT, RSC, RGFA, TML, AT and OCFJ contributed to a detailed revision of the manuscript.

## Acknowledgements

Authors gratefully acknowledge Dr. B. Graham and Dr. K. Corbett (VRC/NIAID/NIH, USA) for sharing the plasmid encoding the S protein gene construct; Prof. E. Durigon (USP, Brazil) for providing the SARS-COV-2 isolate used in neutralization assays; Dr. S. Garcia and Dr. L. Amorim from Hemorio for plasma samples; all study participants for donation of blood samples; and the team of volunteers working in the UFRJ COVID-19 screening center. We thank D. Mucida, C. Lucas and M. Bellio for critical reading and editing of the manuscript. This work was supported by Senai CETIQT, Senai DN and CTG, and by the Brazilian research funding agencies Fundação de Amparo à Pesquisa do Estado do Rio de Janeiro (FAPERJ), Conselho Nacional de Desenvolvimento Científico e Tecnológico (CNPq), Coordenação de Aperfeiçoamento de Pessoal de Nível Superior (CAPES) and Instituto Serrapilheira. DASR was supported by a fellowship from CNPq (DTI-A; 401209/2020-2).

## Supplementary material

**Supplementary Figure S1:**
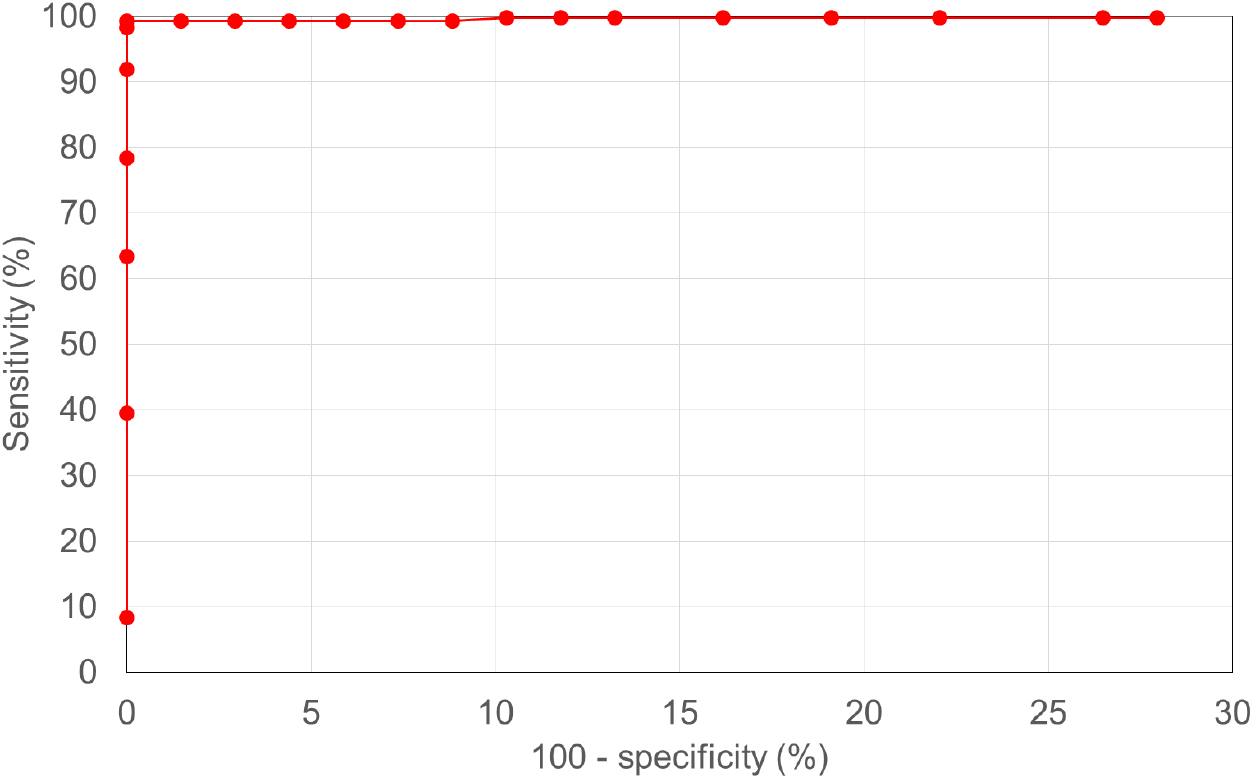
Receiver operating characteristic (ROC) curve of S-UFRJ test. The curve is based on calculating the sensitivity and specificity as a function of varying cut-off values (each data point represents one cut-off value). Sensitivity and specificity were determined based on a panel comprising 420 positive and 68 negative samples.

**Supplementary Table S1:** Cost estimate of consumables needed for S-UFRJ test, considering two scenarios: (1) purchase of materials sold by international suppliers in the country and with no saving efforts; (2) purchase of materials from domestic suppliers and making saving efforts that were validated as not interfering in test performance (data not shown): use of skim milk (2% m/v) instead of BSA (1% m/v); in-house preparation of PBS; finger prick using just the sterile lancet, with no lancing device; and reuse of buffer reservoirs for an equivalent of 30 ELISA plates. As a conservative estimate to account for controls included in each plate and for any eventual repetition needs or failures, a total of 80 tests were considered per each ELISA plate. All quotes for consumables in this table were obtained in June-July 2020 for purchase in the local market in Brazilian reais (BRL). The conversion to USD was done considering an exchange rate of 1 USD = 5.34 BRL (as of July 16, 2020).

| Consumables | Supplier | Amount used (per 80 samples or per plate) | Unit cost (converted to USD) | Package size | Cost per plate (USD) |
| --- | --- | --- | --- | --- | --- |
| Scenario 1 (international suppliers) |  |  |  |  |  |
| Dilution plate | Sarstedt, #821581 (Germany) | 1 | 63.67 | per 100 | 0.64 |
| ELISA plate | Corning, #3590 (USA) | 1 | 146.07 | per 100 | 1.46 |
| S protein | Produced at UFRJ | 20 µg | 140.45 | per mg | 2.81 |
| PBS 10x | Sigma, #11666789001-4L (USA) | 350 mL after 1:10 dilution | 166.67 | per 4000 mL | 1.46 |
| BSA (at 1% m/v) | Sigma, #A2153-100G (USA) | 0.35 g BSA | 344.57 | per 100 g | 1.21 |
| HRP-conjugated antibody | Sigma, #SAB3701282-2MG (USA) | 5 mL after 1:8000 dilution | 261.61 | per mL @ 2 mg/mL | 0.16 |
| TMB | Thermo Fisher, #002023 (USA) | 5 mL | 186.30 | per 500 mL | 1.86 |
| HCl 37% | Sigma, #30721-2.5L (USA) | 5 mL after 1:10 dilution | 32.58 | per 2500 mL | 0.01 |
| Pipette tips | Axygen (USA) | 400 yellow tips | 176.03 | per 20000 | 3.52 |
| Reagent reservoir | Corning Costar (USA) | 6 reservoirs for 30 plates | 146.07 | per 1000 | 0.88 |
| Plastic strips w/ 3 pads | Organicoat (Brazil) | 80 | 243.45 | per 5000 | 3.90 |
| Lancet | G-Tech (Brazil) | 80 | 1.22 | per 100 | 0.97 |
| Lancing device | G-Tech (Brazil) | 1 | 2.79 | per 1 | 2.79 |
| Personal protective equipment (PPE) | Several suppliers | All PPEs (sample collection/analysis) | 2.50 | set of PPEs | 2.50 |
| Additional minor non-listed consumables |  | 20% of all previous items together | 4.83 |  | 4.83 |
|  |  |  | Total costs per plate: |  | 28.99 |
|  |  |  | Cost per test: |  | 0.36 |
| Scenario 2 (domestic suppliers, skim milk, in-house PBS, reservoir reuse, no lanceting device) |  |  |  |  |  |
| Dilution plate | Alfa (Brazil) | 1 | 28.09 | per 50 | 0.56 |
| ELISA plate | Corning, #3590 (USA) | 1 | 146.07 | per 100 | 1.46 |
| S protein | Produced in-house | 20 µg | 140.45 | per mg | 2.81 |
| PBS (1x) (in-house prep) | Synth Chemicals (Brazil) | 350 mL | 0.09 | per liter | 0.03 |
| Skim milk (at 2% m/v) | Nestlé (Switzerland) | 0.7 g milk | 3.04 | per 280 g | 0.01 |
| HRP-conjugated polyclonal antibody | Rhea Biotech (Brazil) | 5 mL after 1:20000 dilution | 196.63 | per mL | 0.05 |
| TMB | Scienco Biotech (Brazil) | 5 mL | 209.74 | per 1000 mL | 1.05 |
| HCl 37% | Sigma, #30721-2.5L (USA) | 5 mL after 1:10 dilution | 32.58 | per 2500 mL | 0.01 |
| Pipette tips | Olen-Econolab (Brazil) | 400 yellow tips | 3.75 | per 1000 | 1.50 |
| Reagent reservoir | Corning Costar (USA) | 6 reservoirs for 30 plates | 146.07 | per 1000 | 0.03 |
| Plastic strips w/ 3 pads | Organicoat (Brazil) | 80 per plate | 243.45 | per 5000 | 3.90 |
| Lancet | G-Tech (Brazil) | 80 per plate | 1.22 | per 100 | 0.97 |
| Personal protective equipment (PPE) | Several suppliers | All PPEs (sample collection/analysis) | 2.50 | set of PPEs | 2.50 |
| Additional minor non-listed consumables |  | 20% of all previous items together | 2.97 |  | 2.97 |
|  |  |  | Total cost per plate: |  | 17.85 |
|  |  |  | Cost per test: |  | 0.22 |

**Supplementary Table S2:** Estimate of the final S-UFRJ test price taking 1 million tests as a calculation basis, including in the cost computation the raw materials shown in Table S1, other manufacturing costs (that include labor, utilities, laboratory charges, insurance, among others) and general expenses (which account for administrative, research, development, distribution and marketing costs)^7^. Profit margin and taxes were also accounted for to estimate the final S-UFRJ test price, since large-scale use of the test for epidemiological surveillance would probably require its industrial production.

| Type of cost | Calculation | Scenario 1 (USD) | Scenario 2 (USD) |
| --- | --- | --- | --- |
| *Raw materials (RM) for 1 million tests* | *See Table S1* | 220000 | 360000 |
| *Other manufacturing costs for 1 million tests* | *145% of RM* | 319000 | 522000 |
| *General expenses for 1 million tests* | *20% of TPC* | 134750 | 220500 |
| Total product costs (TPC) for 1 million tests |  | 673750 | 1102500 |
| Profit margin (PR) (10% of TPC) |  | 67375 | 110250 |
| Taxes (35% of PR) |  | 23581 | 38588 |
| Sum of TPC, PR and taxes |  | 764706 | 1251338 |
| Estimated unit price per test |  | 0.76 | 1.25 |
| Final estimated price per test (average of the 2 scenarios) |  | 1.01 |  |

